# Computational Phenotyping of Electroconvulsive Therapy Outcomes in Treatment-Resistant Depression

**DOI:** 10.1101/2024.10.02.24314373

**Authors:** Rachel E. Jones, L. Paul Sands, Jonathan D. Trattner, Angela Jiang, Christina K. Johnson, Predrag V. Gligorovic, Heather E. Douglas, Rommel Ramos, Kenneth T. Kishida

**Author notes:** **Corresponding Author:** Kenneth T. Kishida, PhD; Wake Forest University School of Medicine, Medical Center Blvd, Winston-Salem NC, 27101, US.

## Abstract

**IMPORTANCE:** Electroconvulsive therapy (ECT) is an effective medical procedure for patients with treatment-resistant depression. However, quantitative neural and behavioral measures that characterize how patients respond to ECT treatment are largely lacking.

**OBJECTIVE:** Determine whether neurocomputational models that integrate information about adaptive learning behavior and associated affective experiences can characterize neurobehavioral changes in patients whose depression improves following ECT treatment.

**DESIGN:** This observational study included two research visits from 2020-2023 that occurred before and after standard-of-care ECT for treatment-resistant depression. This report focuses on “visit 2”, which occurred after patients received their initial ECT treatment series.

**SETTING:** Wake Forest University School of Medicine; Atrium Health Wake Forest Baptist Psychiatric Outpatient Center; Atrium Health Wake Forest Hospital.

**PARTICIPANTS:** Participants who received ECT for treatment-resistant depression (“ECT”), and participants not receiving ECT but with depression (“non-ECT”) or without depression (“no-depression”) were recruited from the Psychiatric Outpatient Center and community, respectively.

**EXPOSURES:** Computerized delivery of a Probabilistic Reward and Punishment with Subjective Rating task with functional magnetic resonance imaging.

**MAIN OUTCOMES AND MEASURES:** Computational modeling of choice behavior provided parameters that characterized learning dynamics and associated affect dynamics expressed through intermittent Likert scale self-reports. Multivariate statistical analyses relating model parameters, neurobehavioral responses, and clinical assessments.

**RESULTS:** ECT (N=21; 47.6% female), non-ECT (N=36; 69.4% female), and no-depression (N=38; 65.8% female) participants. Parameters derived from computational models fit to behavior elicited during learning *and* the expression of affective experiences for all groups reveled specific changes in patients who responded favorably to ECT. ECT-responders demonstrated increased rates of learning from rewarding trials, normalized affective response to punishments, and an increase in the influence of counterfactual ‘missed opportunities’ on affective behavior. Additionally, ECT-responders’ showed changes in BOLD activity regions specific to each of these parameters. ECT-responders’ BOLD-responses to surprising punishments and counterfactual missed opportunities were altered from visit 1 to visit 2 in the inferior frontal operculum, Rolandic operculum, precentral gyrus, and caudate.

**CONCLUSIONS AND RELEVANCE:** Computational models of neurobehavioral dynamics associated with learning and affect can describe specific hypotheses about neurocomputational-mechanisms underlying favorable responses to treatment-resistant depression. Our results suggest computational estimates of learning and affective dynamics may aid in identifying depression phenotypes and treatment outcomes in psychiatric medicine where objective measures are largely lacking.

**KEY POINTS:** *QUESTION:* How does ECT treatment change neurocomputational measures of learning and affective experiences in patients with treatment-resistant depression?

*FINDINGS:* In this observational study, computational models were used to quantify the behavioral dynamics of learning and associated changes in subjective feelings in patients who underwent ECT treatment for treatment resistant depression and controls. In ECT-responders we observed increases in reward-based learning, normalized affective responses to surprising positive and negative outcomes, and associated changes in fMRI-measured BOLD-responses.

*MEANING:* Computational phenotyping of task behavior and associated brain responses provides quantification of complex neurobehavioral dynamics and provides specific insight into the neurobehavioral mechanisms underlying successful ECT treatment.

## INTRODUCTION

Electroconvulsive therapy (ECT) is an effective medical procedure for patients with treatment-resistant depression (TRD) who are unresponsive to standard antidepressants^1,2^. Despite decades of use, it is still unclear how ECT-induced changes in brain function give rise to behavioral changes in treatment responders^3–5^. Objective measures that link neural and behavioral changes produced by ECT treatment may provide a precise explanation of how ECT improves depressive symptoms in patients with TRD^6–8^.

Computational models of reinforcement learning (RL) have been increasingly applied to better understand depression pathophysiology and treatment mechanisms^9–11^. Further, quantifying links between measurable and computable learning signals and subjective emotional states has started to reveal how depression impacts the emotional processing of decision outcomes^12–15^. However, to date, no study has applied neurocomputational depictions of learning and affective experiences to characterize ECT treatment outcomes. The goal of this study was to combine computational models of learning and affective dynamics with functional magnetic resonance imaging (fMRI) to characterize neurobehavioral changes in patients with TRD following their initial ECT treatment series. Specifically, we used a Valence Partitioned Reinforcement Learning (VPRL) model recently shown to explain human choice behavior better than single-valence RL models and that was shown to explain sub-second changes in dopamine fluctuations in humans performing the same task that is used in this work^16–18^.

While computational RL methods describing post-ECT effects are lacking, other clinical treatments have been shown to improve reward learning deficits in non-treatment resistant depression^19–24^. For example, antidepressants increase neural responses to reward prediction errors in responsive patients^25–27^. However, there are variable findings regarding punishment learning^28–30^. For example, multiple fMRI studies using similar tasks report inconsistent BOLD responses to ‘negative reward prediction errors’ in unmedicated patients with depression^31–35^. Further, antidepressant studies also report increased^36^, decreased^37^, or no change^38,39^ in patients’ punishment learning when considering ‘negative reward prediction errors’ and the punishment signal.

Brown and colleagues recently separated trials within their RL tasks according to whether the trial was rewarding or punishing (e.g., gains vs. losses, respectively) and demonstrated normalized reward and loss neurocomputations after cognitive behavioral therapy in patients with depression^9^. This separation of valence aligns with recent evidence suggesting positive and negative valence processing occurs through separate neural systems, which differs from traditional single-valence RL approaches^28,40–42^. The VPRL framework used in this study follows this line of reasoning and hypothesizes that independent neural systems track positive and negative events whether anticipated or actually experienced^16–18,28^. We hypothesize that applying VPRL to investigate ECT treatment outcomes may clarify how ECT alters distinct positive and negative learning mechanisms in patients with TRD.

In addition to reinforcement learning and decision making mechanisms, further investigating how positive and negative learning signals differentially influence affective states using neurocomputational models, may identify specific affective mechanisms altered by ECT treatment^43–48^. For example, Eldar and Niv demonstrated that positive prediction errors improved mood while negative prediction errors worsened mood during a learning task, with emotional states further biasing valuations of subsequent outcomes in individuals with mood instabilities^13^. Also, Rutledge and colleagues showed that reward expectations and reward prediction errors directly affected mood ratings about recent outcomes in patients with depression^12^. While such studies dig deeper into the influence of decision outcomes on affective state, there remains a gap in the literature describing how neurobehavioral computations may give rise to affective experiences and how these processes are altered in depression and perhaps favorably modulated by effective treatments including ECT^49,50^.

In this study, we applied a VPRL framework^17,18^ to determine whether a computational psychiatric approach^51,52^ that uses valenced-partitioned (i.e., positive and negative systems) learning signals to predict affective behavior could provide insight into how ECT treatment changes learning and affective dynamics in patients with favorable responses. We hypothesized that ECT would alter neurobehavioral signals related to both learning and affective behavior in ECT treatment responders.

## METHODS

### Study Design

Here, we report data collected during research “visit 2” of a two-visit observational study following a standard-of-care ECT-treatment timeline (Fig.1A). All participants completed a Probabilistic Reward and Punishment with Subjective Rating task during fMRI scanning. Following fMRI scanning, participants completed the Patient Health Questionnaire 9 (PHQ-9)^53^, Hamilton Depression Rating Scale (HAM-D)^54^, and Montreal Cognitive Assessment (MOCA)^55^. Participants provided written informed consent under Wake Forest University School of Medicine IRB00056131.

**Figure 1.**
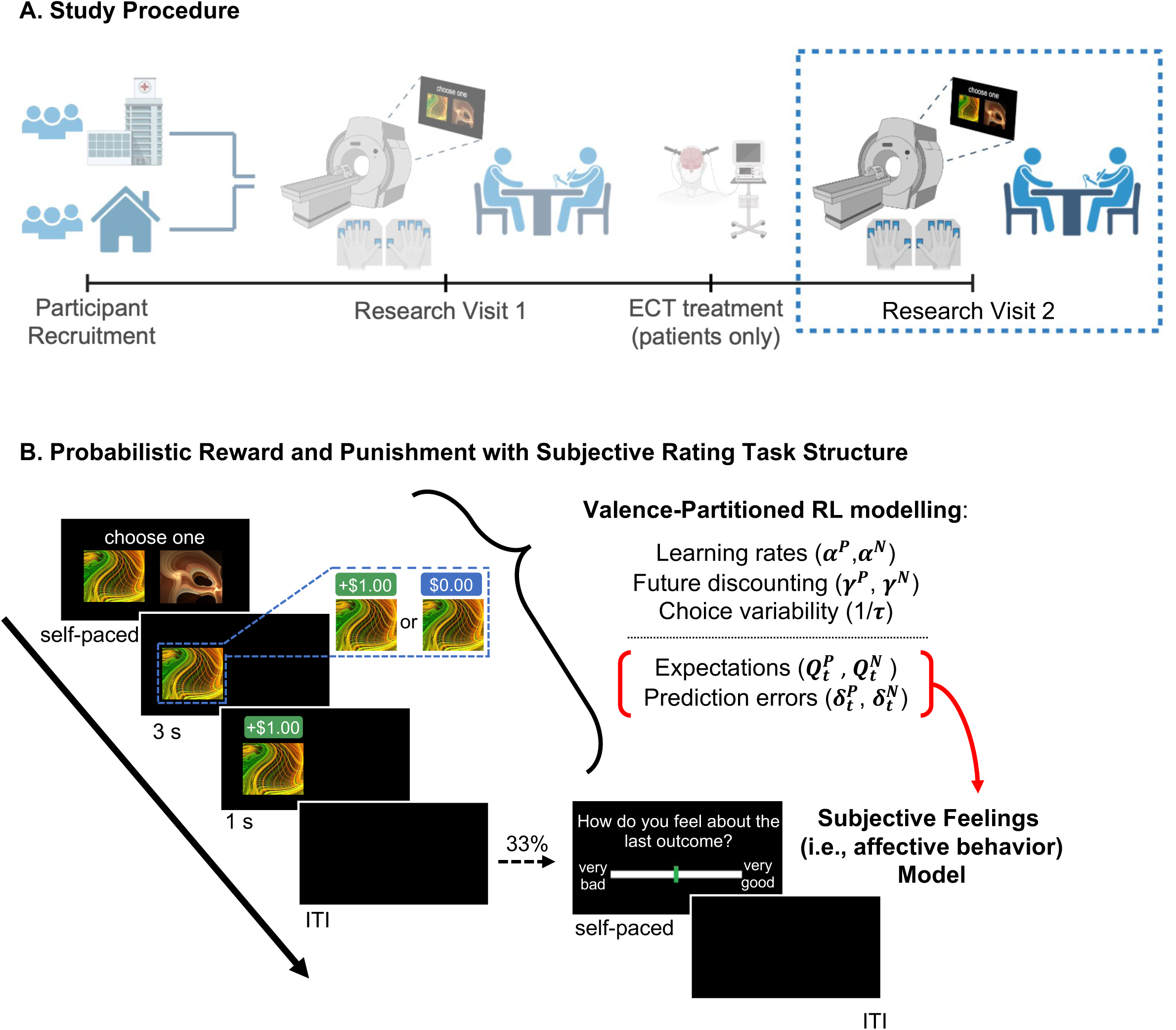
Study design. **(A)** This study focuses on research visit 2 results, which took place after patients with treatment-resistant depression received ECT treatment for the first time and approximately one to two months following visit 1 for non-ECT and non-depression participants (to follow along the ECT treatment timeline). All participants completed a probabilistic reward and punishment with subjective rating (PRPwSR) task while receiving fMRI scanning and completed clinical assessments afterward; these measures were identical to those taken at visit 1. **(B)** In the PRPwSR task, each trial starts with an ‘option presentation’ screen. A participant chooses an option (self-paced) and then the other option disappears. After 3s, the chosen option is reinforced probabilistically (monetary gain, no gain, or loss), and the monetary outcome is shown for 1s. The screen then goes blank for a random-length interval or displays with 33% probability a subjective rating screen (self-paced) followed by the blank screen before the next trial. We fit a valence-partitioned reinforcement learning model to participants’ choice behavior in the PRPwSR task to generate learning parameters. Expectations and prediction errors (depicted in red brackets) were then fitted as independent predictors of participant-reported subjective rating in a regression model (i.e., Subjective Feeling Model).

### Participants

“ECT” patients included patients with TRD who received ECT treatment for the first time at AHWFB Psychiatric Outpatient Center (ECT, N=21; 47.6% female). We defined ECT treatment responders as patients who showed any clinical improvement following their standard-of-care ECT treatment series based on clinician notes or PHQ-9 or HAM-D assessments (ECT Responder, N=17; ECT Non-responder, N=4). Participants with depression not planning ECT (non-ECT, N=36; 69.4% female) and participants without depression (no-depression, N=38; 65.8% female) were recruited from the Winston-Salem, North Carolina area. See eMethods and eTables1-3 for full participant details.

### Probabilistic Reward and Punishment with Subjective Rating (PRPwSR) Task

Participants completed the same PRPwSR task completed at research visit 1 (Fig.1B)^17,18^. Briefly, the PRPwSR task is a value-based choice task where participants make choices and learn to maximize probabilistic rewards (i.e., monetary gains) and minimize probabilistic losses (i.e., monetary losses) over 150 trials. After each trial there is a one-third probability that participants would be asked, “How do you feel about the last outcome?”. Participants respond using a Likert scale ranging from “very bad” to “very good.” See eMethods and eFig.1 for task details. See eFig.2 for group performance measures.

### Computational Modeling

We used hierarchical Bayesian methods^56^ to fit RL computational models to participants’ choice behavior on the PRPwSR task (eMethods). Participants’ expected icon values and outcome prediction errors were estimated using a VPRL framework^16–18^. These learning signals were then used to model and predict participants’ subjective ratings during the task. We hypothesize that each individual’s unique set of learning and affective parameters could represent a *computational phenotype* and that ECT responders’ computational phenotype would change relative to pre-ECT-treatment. See below for brief descriptions of model parameters:

### Valence Partitioned Reinforcement Learning Model Parameters

A previously validated VPRL framework was used to quantify participants’ choice behavior^16–18^. VPRL hypothesizes that brains track appetitive (e.g., positive/rewarding) and aversive (e.g., negative/punishing) stimuli simultaneously, but via independent systems. This allows stimuli to predict benefits and/or costs that may be independently estimated such that cost-benefit comparisons can be made^16^. Each system updates *value estimates* (i.e., Q-values) akin to standard Q-learning with temporal difference RL rules^57^. Hence, Q-values for positive 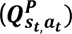 and negative 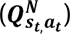 state-action sets (*s_t_*, *a_t_*) are estimated. Likewise, future states’ (*S_t_*_+1_) positive 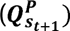 and negative 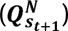 values are estimated and, respectively, discounted by independent parameters (γ^P^,γ^N^). The combination of current actual outcomes, discounted expectations of future values, and expected current values are used to calculate independent temporal difference prediction errors 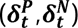, which are then used to update respective Q-values by independent learning weights (*α^P^*, *α^N^*). We modeled choice policy using a softmax function with a temperature parameter (*τ*) random a participant’s choices are given Q-value estimates. The parameters in the VPRL framework (*α^P^*, *α^N^*, *γ^P^*, *γ^N^*, and *τ*) were used to characterize each participant’s computational *learning* phenotype. See eMethods for details about the VPRL model.

### Subjective Feeling Regression Model Parameters

We hypothesized that participants’ expectations about icon values 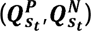 and prediction errors resulting from choice outcomes 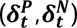 collectively contributed to their reported feelings in the PRPwSR task^18^. We also hypothesized that the contribution of learning signals to subjective ratings in ECT-responders would differ from those observed before ECT treatment. We fit a linear regression model^58^ with Q-values 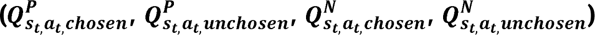 and prediction errors 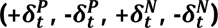 as independent predictors of subjective ratings. The coefficient parameters in the Subjective Feeling model 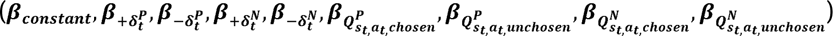 were used to characterize each participant’s computational *affective* phenotype. See eMethods for details about the Subjective Feeling model.

### fMRI Analyses

See eMethods for details on fMRI data acquisition, pre-processing, and model-based analyses. VPRL and Subjective Feeling models were fit to each participants’ behavior. We developed second-level “visit 2” minus “visit 1” contrasts of specific learning and affective neurocomputations that showed behavioral changes across research visits and performed whole-brain i) one-sample t-tests of blood-oxygen-level-dependent (BOLD) changes for ECT responders and ii) analysis of the variance (ANOVA) of BOLD changes between ECT, non-ECT, and no-depression groups. All statistical analyses were conducted at an uncorrected threshold of p<0.001 and reported results were selected using a family-wise error (FWE)-corrected threshold of p<0.05 at cluster and peak voxel levels.

### Statistical Analysis

We used hierarchical Bayesian analysis^56^ to estimate posterior distributions of free parameters in the VPRL model (*α^P^*, *α^N^*, *γ^P^*, *γ^N^*, and *τ*) and subsequently conducted a Bayesian linear regression^58^ with group-informed posterior coefficients in the Subjective Feeling model 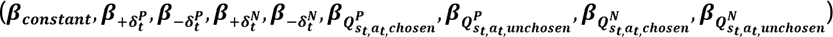. For all relevant tests, p<0.05 was deemed significant. Analyses were conducted in R version 4.2.2 and Stan version 2.21.0. (rstan version 2.21.8)^59^.

## RESULTS

### Participant characteristics

Across research visits, both ECT and non-ECT groups demonstrated decreases in PHQ-9 and HAM-D scores, indicating depression symptom improvement. ECT patients showed a substantial reduction in PHQ-9 scores compared to both non-ECT and no-depression groups but a substantial reduction in HAM-D scores compared to no-depression participants only. The three groups did not differ in age, gender, race, or ethnicity. See eTable4 for clinical details.

### ECT Responder *Computational Phenotypes* for Valence Partitioned Learning

#### Behavior

We hypothesized that ECT treatment altered neurobehavioral learning mechanisms in responders. To test this hypothesis, we fit the VPRL model separately to ECT responder, ECT non-responder, non-ECT, and no-depression group choice behavior for both research visits and assessed group-level changes in learning parameters across visits.

ECT responders showed an increase in the positive system learning rate (*α^P^*, Table 1A) following treatment (median difference [95% HDI] = 0.15[−0.05, 0.35]). No-depression participants also showed some increase in *α^P^* (0.02 [−0.04, 0.09]) while non-ECT participants showed decreased *α^P^* across research visits (−0.09 [−0.15, −0.02]). Non-ECT and no-depression groups also showed decreases in 1/.t (non-ECT: −0.08 [−0.20, 0.05]; no-depression: −0.04 [−0.13, 0.04]) across visits.

**Table 1.** Group-level changes in learning and subjective experience behavioral computations across research visits.

|  |  |  |  |  |  |  |  |  |  |  |  |  |  |  |  |  |
| --- | --- | --- | --- | --- | --- | --- | --- | --- | --- | --- | --- | --- | --- | --- | --- | --- |
|  | ECT Responders (n = 17) |  |  |  | ECT Non-responders (n = 4) |  |  |  | Non-ECT (n = 34) |  |  |  | No-depression (n = 38) |  |  |  |
|  | Pre-ECT | Post-ECT | Median<br>Δ<br>[95% HDI] | Credible<br>Difference | Pre-ECT | Post-ECT | Median<br>Δ<br>[95% HDI] | Credible<br>Difference | Visit 1 | Visit 2 | Median<br>Δ<br>[95% HDI] | Credible<br>Difference | Visit 1 | Visit 2 | Median<br>Δ<br>[95% HDI] | Credible<br>Difference |
|  | Median<br>[95% HDI] | Median<br>[95% HDI] |  |  | Median<br>[95% HDI] | Median<br>[95% HDI] |  |  | Median<br>[95% HDI] | Median<br>[95% HDI] |  |  | Median<br>[95% HDI] | Median<br>[95% HDI] |  |  |
| A. Learning Parameters |  |  |  |  |  |  |  |  |  |  |  |  |  |  |  |  |
| $\alpha^P$ | 0.15<br>[0.04, 0.31] | 0.31<br>[0.16, 0.46] | **0.15**<br>**[-0.05, 0.35]** | [7.94, 92.06] | 0.12<br>[0.04, 0.29] | 0.22<br>[0.11, 0.38] | 0.10<br>[-0.12, 0.30] | [13.63, 86.38] | 0.21<br>[0.15, 0.27] | 0.12<br>[0.08, 0.15] | **-0.09**<br>**[-0.15, -0.02]** | [99.79, 0.21] | 0.16<br>[0.13, 0.20] | 0.18<br>[0.13, 0.24] | **0.02**<br>**[-0.04, 0.09]** | 24.82, 75.17 |
| $\gamma^P$ | 0.70<br>[0.41, 1.0] | 0.54<br>[0.32, 0.84] | -0.15<br>[-0.56, 0.26] | [74.74, 25.26] | 0.72<br>[0.44, 1.0] | 0.81<br>[0.53, 1.0] | 0.08<br>[-0.35, 0.48] | [36.18, 63.82] | 0.83<br>[0.51, 1.0] | 0.79<br>[0.60, 1.0] | -0.02<br>[-0.35, 0.39] | [54.17, 45.82] | 0.83<br>[0.56, 1.0] | 0.73<br>[0.55, 0.97] | -0.07<br>[-0.38, 0.29] | 64.45, 35.55 |
| $\alpha^N$ | 0.45<br>[0.02, 0.90] | 0.25<br>[4.35e<sup>-4</sup>, 0.63] | -0.19<br>[-0.83, 0.40] | [70.34, 29.66] | 0.28<br>[1.48e<sup>-5</sup>, 7.34e<sup>-1</sup>] | 0.42<br>[1.40e<sup>-5</sup>, 0.93] | 0.11<br>[-0.56, 0.86] | [38.70, 61.30] | 0.46<br>[3.2e<sup>-4</sup>, 0.65] | 0.52<br>[0.08, 0.78] | 0.07<br>[-0.42, 0.62] | [38.20, 61.80] | 0.53<br>[0.02, 0.76] | 0.41<br>[0.03, 0.73] | -0.08<br>[-0.58, 0.53] | 60.06, 39.94 |
| $\gamma^N$ | 0.23<br>[8.64e<sup>-3</sup>, 0.53] | 0.25<br>[3.68e<sup>-3</sup>, 0.81] | 0.03<br>[-0.42, 0.70] | [44.24, 55.76] | 0.34<br>[1.77e<sup>-3</sup>, 0.89] | 0.15<br>[1.07e<sup>-5</sup>, 0.68] | -0.15<br>[-0.86, 0.47] | [69.94, 30.06] | 0.11<br>[1.0e<sup>-4</sup>, 0.58] | 0.15<br>[0.02, 0.38] | 0.04<br>[-0.50, 0.39] | [38.41, 61.59] | 0.19<br>[0.02, 0.49] | 0.23<br>[0.04, 0.51] | 0.03<br>[-0.35, 0.41] | 43.08, 56.93 |
| 1/τ | 0.10<br>[0.05, 0.27] | 0.08<br>[0.05, 0.17] | -0.01<br>[-0.21, 0.12] | [61.62, 38.38] | 0.08<br>[0.05, 0.17] | 0.08<br>[0.05, 0.15] | -6.17e<sup>-4</sup><br>[-0.13, 0.10] | [50.86, 49.14] | 0.17<br>[0.05, 0.26] | 0.08<br>[0.05, 0.13] | **-0.08**<br>**[-0.20, 0.05]** | [85.47, 14.52] | 0.11<br>[0.05, 0.19] | 0.07<br>[0.05, 0.11] | **-0.04**<br>**[-0.13, 0.04]** | 82.35, 17.65 |
| B. Subjective Experience Parameters |  |  |  |  |  |  |  |  |  |  |  |  |  |  |  |  |
| $\beta_{+\delta_t^P}$ | 0.83<br>[0.67, 1.0] | 0.70<br>[0.54, 0.85] | -0.14<br>[-0.36, 0.09] | [87.44, 12.56] | 0.88<br>[0.44, 1.28] | 1.01<br>[0.44, 1.42] | 0.14<br>[-0.62, 0.74] | [35.30, 64.70] | 0.83<br>[0.73, 0.93] | 0.91<br>[0.81, 1.0] | 0.08<br>[-0.06, 0.22] | [13.97, 86.03] | 0.90<br>[0.77, 1.04] | 0.88<br>[0.75, 1.02] | -0.02<br>[-0.21, 0.17] | 58.34, 41.66 |
| $\beta_{-\delta_t^P}$ | -0.39<br>[-0.69, -0.10] | -0.49<br>[-0.71, -0.28] | -0.10<br>[-0.47, 0.26] | [70.81, 29.19] | 0.18<br>[-0.61, 0.95] | -0.20<br>[-0.74, 0.33] | -0.36<br>[-1.40, 0.62] | [77.42, 22.58] | -0.44<br>[-0.61, -0.28] | -0.39<br>[-0.57, -0.20] | 0.06<br>[-0.19, 0.31] | [32.29, 67.71] | -0.03<br>[-0.28, 0.22] | -0.61<br>[-0.84, -0.38] | **-0.58**<br>**[-0.93, -0.24]** | 99.96, 0.04 |
| $\beta_{Q_{s_t, a_t, chosen}^P}$ | 1.10<br>[0.59, 1.62] | 0.96<br>[0.44, 1.47] | -0.14<br>[-0.86, 0.62] | [64.45, 35.55] | 1.25<br>[-0.16, 2.57] | 0.61<br>[-0.34, 1.70] | -0.63<br>[-2.34, 1.28] | [74.80, 25.21] | 1.11<br>[0.83, 1.38] | 0.97<br>[0.64, 1.30] | -0.14<br>[-0.56, 0.29] | [73.27, 26.73] | 0.87<br>[0.49, 1.25] | 1.89<br>[1.43, 2.34] | **1.01**<br>**[0.40, 1.59]** | 0.10, 99.90 |
| $\beta_{Q_{s_t, a_t, unchosen}^P}$ | -0.61<br>[-1.60, 0.34] | 0.65<br>[-0.28, 1.59] | **1.27**<br>**[-0.04, 2.64]** | [3.14, 96.86] | 0.22<br>[-1.41, 1.89] | 0.31<br>[-1.14, 1.66] | 0.07<br>[-2.02, 2.24] | [47.33, 52.68] | -0.58<br>[-0.97, -0.19] | 0.26<br>[-0.36, 0.89] | **0.84**<br>**[0.10, 1.58]** | [1.30, 98.70] | 0.33<br>[-0.35, 1.03] | 0.75<br>[-0.07, 1.59] | 0.42<br>[-0.67, 1.49] | [22.58, 77.42] |

|  | *ECT Responders (n = 17) (continued)* |  |  |  | *ECT Non-responders (n = 4) (continued)* |  |  |  | *Non-ECT (n = 34) (continued)* |  |  |  | *No-depression (n = 38) (continued)* |  |  |  |
| --- | --- | --- | --- | --- | --- | --- | --- | --- | --- | --- | --- | --- | --- | --- | --- | --- |
| | Pre-ECT | Post-ECT | Median<br>$\Delta$<br>[95% HDI] | Credible<br>Difference | Pre-ECT | Post-ECT | Median<br>$\Delta$<br>[95% HDI] | Credible<br>Difference | Visit 1 | Visit 2 | Median<br>$\Delta$<br>[95% HDI] | Credible<br>Difference | Visit 1 | Visit 2 | Median<br>$\Delta$<br>[95% HDI] | Credible<br>Difference |
|  | Median<br>[95% HDI] | Median<br>[95% HDI] |  |  | Median<br>[95% HDI] | Median<br>[95% HDI] |  |  | Median<br>[95% HDI] | Median<br>[95% HDI] |  |  | Median<br>[95% HDI] | Median<br>[95% HDI] |  |  |
| *B. Subjective Experience Parameters (continued)* |  |  |  |  |  |  |  |  |  |  |  |  |  |  |  |  |
| $\beta_{+\delta_t^N}$ | -0.67<br>[-0.97, -0.38] | -1.32<br>[-1.65, -1.0] | **-0.65**<br>**[-1.09, -0.22]** | [99.83, 0.17] | -1.10<br>[-1.73, -0.52] | -0.92<br>[-1.65, -0.20] | 0.18<br>[-0.82, 1.23] | [36.92, 63.08] | -1.23<br>[-1.40, -1.04] | -1.11<br>[-1.29, -0.92] | 0.12<br>[-0.14, 0.38] | [17.76, 82.24] | -1.08<br>[-1.33, -0.82] | -0.98<br>[-1.23, -0.72] | 0.10<br>[-0.26, 0.46] | [28.51, 71.49] |
| $\beta_{-\delta_t^N}$ | 0.19<br>[-0.18, 0.56] | -0.38<br>[-0.82, 0.05] | **-0.57**<br>**[-1.14, -2.76e<sup>-4</sup>]** | [97.39, 2.61] | 0.18<br>[-0.84, 1.23] | -0.48<br>[-1.40, 0.43] | -0.66<br>[-1.96, 0.61] | [84.34, 15.66] | 0.15<br>[-0.12, 0.41] | -0.05<br>[-0.19, 0.30] | -0.09<br>[-0.45, 0.27] | [69.49, 30.51] | 0.24<br>[-0.08, 0.56] | -0.34<br>[-0.69, 8.6e<sup>-3</sup>] | **-0.58**<br>**[-1.06, -0.10]** | [99.13, 0.87] |
| $\beta_{Q_{s_t, a_t, chosen}^N}$ | -0.37<br>[-2.07, 1.30] | -0.56<br>[-2.07, 0.99] | -0.19<br>[-2.49, 2.09] | [56.45, 43.55] | -0.76<br>[-2.56, 1.33] | -0.21<br>[-2.12, 1.77] | 0.53<br>[-2.17, 3.31] | [35.26, 64.74] | -1.68<br>[-2.96, -0.36] | -1.67<br>[-2.98, -0.36] | 0.01<br>[-1.83, 1.85] | [49.49, 50.51] | -1.39<br>[-3.02, 0.22] | -0.68<br>[-2.11, 0.68] | 0.71<br>[-1.40, 2.85] | [25.51, 74.49] |
| $\beta_{Q_{s_t, a_t, unchosen}^N}$ | -0.68<br>[-2.12, 0.73] | 0.54<br>[-0.45, 1.54] | 1.22<br>[-0.46, 2.98] | [8.39, 91.61] | -0.13<br>[-1.64, 1.39] | 0.04<br>[-1.81, 1.99] | 0.15<br>[-2.26, 2.56] | [44.97, 55.03] | 0.29<br>[-0.48, 0.20] | 0.40<br>[-0.54, 1.36] | 0.11<br>[-1.07, 1.29] | [42.69, 57.31] | 1.80<br>[0.63, 2.97] | 1.08<br>[0.13, 2.02] | -0.71<br>[-2.16, 0.82] | [82.79, 17.21] |
| Bold indicates evidence of distributional differences. Abbreviations: HDI: highest density interval. |  |  |  |  |  |  |  |  |  |  |  |  |  |  |  |  |

#### Neural

Given the change in *α^P^* among ECT responders, we performed a one-sample t-test of BOLD change across research visits (visit 2-visit 1) associated with positive system reward prediction errors 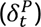 in ECT responders (see Supplementary Eq.1-2). However, we did not find significant changes in this activity following treatment (Table 2A). We then tested whether BOLD activity associated with 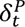 changed more in the ECT cohort compared to non-ECT and no-depression groups. We did not find significant differences between the three cohorts after conducting a whole-brain ANOVA on visit 2-visit 1 change in BOLD-associated 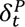 activity (eTable5A).

**Table 2.** Paired t-test for ECT responder changes in learning and subjective experience neurocomputations following treatment.

|  |  |  |  |  |  |  |
| --- | --- | --- | --- | --- | --- | --- |
| Parameter | Region | Voxels | Cluster-level p-value | Peak-level p-value | T-statistic | Peak MNI coordinates [x y z] |
| *A. Learning Computations* |  |  |  |  |  |  |
| $\Delta_{visit\ 2-visit\ 1}(\beta_{GLM}(\delta_t^P))$ | | | | | NS | |
| *B. Subjective Experience Computations* |  |  |  |  |  |  |
| $\Delta_{visit\ 2-visit\ 1}(\beta_{GLM}(\beta_{Q_{st,\alpha_t,unchosen}^P}))$ | Left rolandic operculum | 92 | **0.009** | 0.689 | T(14) = 6.55 | [-40 2 16] |
|  | Left caudate | 144 | **0.001** | 0.921 | T(13) = 5.86 | [-16 24 10] |
| $\Delta_{visit\ 2-visit\ 1}(\beta_{GLM}(\beta_{+\delta_t^N}))$ | Right inferior frontal operculum | 279 | **7.78e-7** | 0.366 | T(13) = 7.37 | [40 8 28] |
| $\Delta_{visit\ 2-visit\ 1}(\beta_{GLM}(\beta_{-\delta_t^N}))$ | Right precentral gyrus | 79 | **0.012** | **0.032** | T(13) = 9.19 | [42 -20 58] |
| All analyses were performed at an uncorrected threshold of p<0.001 in which reported results were selected using an FWE-corrected threshold of p<0.05 at cluster and peak voxel levels. Bold font indicates significance. |  |  |  |  |  |  |

### ECT Responder *Computational Phenotypes* for Subjective Feelings

#### Behavior

We hypothesized that i) VPRL learning signals may drive changes in subjective feelings about the consequences of choices made (Fig.1B) and ii) ECT treatment would alter this affective mechanism in responders for specific learning signal contributions on reported feelings. We expected a linear combination of these signals: 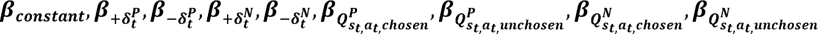 to predict subjective ratings.

In ECT-responders only, receiving worse than expected punishments 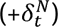 contributed to more negative ratings of feelings after treatment (median difference [95% HDI] = −0.65 [−1.09, −0.22], Table 1B). Both ECT responders and non-ECT participants demonstrated a shift from a negative to positive influence of positive system counterfactual choice expectations 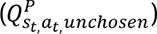 on feelings across visits (ECT responders: 1.27 [−0.04, 2.64]; non-ECT: 0.84 [0.10, 1.58]). Alternatively, both ECT responders and no-depression participants shifted from a positive to negative influence of ‘less-bad than expected’ punishments 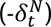 on rated feelings (ECT responders: −0.57 [−1.14, −2.76e-4]; no-depression: −0.58 [−1.06, −0.10]). No-depression participants demonstrated more negative feelings derived from less rewarding outcomes 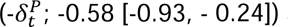 and more positive feelings from positive system chosen choice expectations 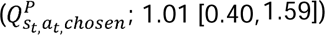.

#### Neural

Given the changes in the Subjective Feeling model parameters we observed in ECT responders, we performed a one-sample t-test to determine BOLD activity change associated with subjective ratings influenced by 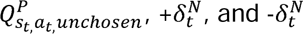 across research visits (visit 2-visit 1). We found BOLD-associated changes in all parameters for ECT responders, with increased activity in the right inferior frontal operculum and decreased activity in the left rolandic operculum, left caudate, and right precentral gyrus following treatment (Fig.2; Table 2B).

**Figure 2.**
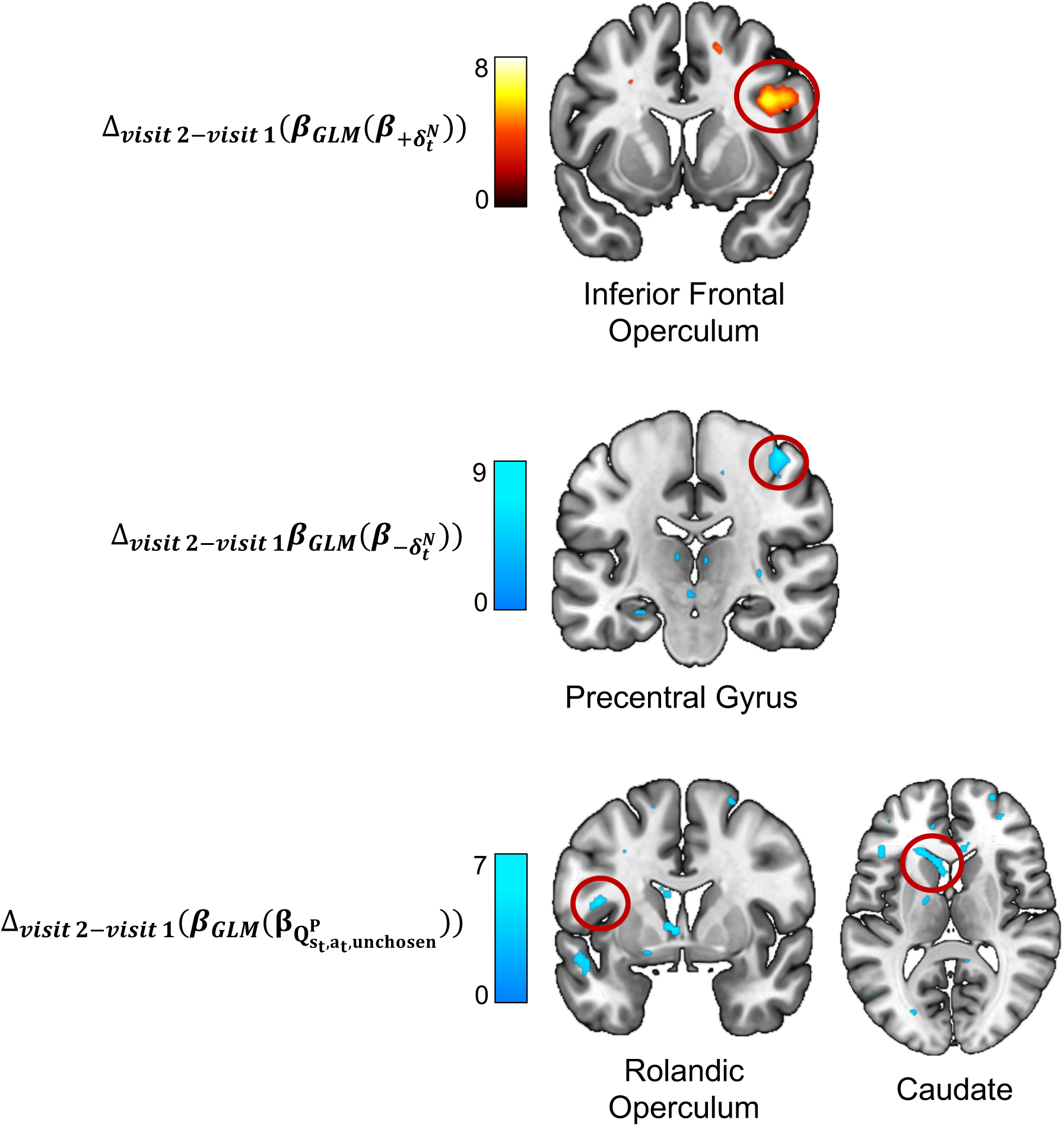
Neural Changes Associated with Subjective Experience Computations in ECT Responders Following Treatment. Between research visits 1 and 2, ECT responders showed increased BOLD activity associated with 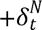 influence on subjective experience (top), and decreased BOLD activity associated with 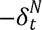 influence on subjective experience (middle) and 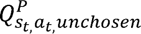 influence on subjective experience (bottom).

To assess whether BOLD-associated changes in subjective feeling computations differed across ECT, non-ECT, and no-depression groups, we performed a whole-brain ANOVA based on behavioral results (we again assessed for group differences in emotional impacts of 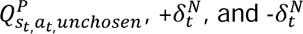. We found differences in BOLD-associated punishment prediction error changes across the groups (eTable5B). Post-hoc t-tests showed greater changes in BOLD-related activity for non-ECT participants compared to ECT patients in the right calcarine gyrus, right precuneus, and left posterior cingulum and for no-depression participants compared to ECT patients in the right angular gyrus.

## DISCUSSION

We investigated whether and how ECT treatment changed computational phenotypic depictions of the neurobehavioral dynamics of reward and punishment learning and associated subjective experiences in patients with TRD (who were previously naïve to ECT). We assessed specific changes in ECT-responders, defined by any clinical improvement in depression, participants with depression managed by medications or therapy, and participants without depression to pinpoint treatment specific effects versus general neurobehavioral changes that may have occurred across research visits. We used a hierarchical Bayesian approach to fit a Valence Partitioned Reinforcement Learning (VPRL)^16–18^ model to participant choice behavior on a Probabilistic Reward and Punishment with Subjective Rating (PRPwSR) task (Fig.1B). This model provided parameters describing learning mechanisms in which ECT responders demonstrated an increase in reward learning rate following treatment (Table 1A). From VPRL parameters, we then estimated participants’ expectations and prediction errors to model their influence on participants’ subjective feelings of decision outcomes during the task. Following treatment, ECT-responders demonstrated unique shifts in how unchosen potentially good (i.e., ‘missed opportunities’) and actual worse-than-expected losses affected their feelings, suggesting a treatment effect in positive and negative RL-affective systems (Table 1B). These specific changes in subjective experience were further associated with changes in BOLD-responses associated with hypothesized affective processes (Fig.2). Our collective results suggest that specific neurobehavioral mechanisms underlying learning and affective processes are altered by successful ECT treatment. Further, these signals may provide a potential quantitative measure for ECT-responders and a favorable ECT response.

To our knowledge, our study is the first to apply a computational psychiatric approach to investigate ECT treatment mechanisms in patients with TRD, and to pair VPRL^16–18^ and related models to extract subjective experience^16,18^ in patients with depression. ECT responders demonstrated increased reward learning rates following treatment compared to other cohorts, suggestive of an ECT treatment effect that may allow relearning of the relationship between potentially good experiences and behaviors that promote them. This is consistent with prior work demonstrating that various types of antidepressants can improve reward learning in responsive patients with depression^19,26,27^, but demonstrates the role that ECT can have in this specific process in treatment resistant patients. Notably, the ECT patients in this study had tried multiple (and were currently taking) antidepressants that were ineffective and therefore proceeded to ECT treatment. These findings suggest ECT may be effective in targeting similar neural circuits associated with reward processing that the mechanisms of antidepressants could not alter for patients with TRD in our study^29,38^. We did not find neural correlates to reward prediction error signaling improvements in ECT treatment responders, indicating further work is needed to pinpoint neural effects of reward learning from ECT.

We found both neural and behavioral changes in how learning signals come to drive affective responses in ECT responders. These results suggest that ECT may have changed mechanisms underlying how good and bad outcomes are processed in patients and how they come to affect subjective feelings. Interestingly, counterfactual ‘missed opportunities’ showed a significant increase in the influence on positive feelings in both ECT-responders and no-depression groups across visits. Originally this parameters promoted strong negative feelings in responders, but after treatment, promoted positive feelings similar to what we observed in no-depression participants. Individuals with depression often experience challenges in experiencing pleasure or motivation (i.e., anhedonia). Despite not seeking rewards, this may cause rumination over missed opportunities and lead to negative feelings^7,60,61^. Our results suggest ECT may normalize this affective mechanism and involve regions such as the caudate and rolandic operculum known to be involved with reward seeking and emotional processing^62,63^. Further, ‘worse-than-expected’ punishments contributed to more negative feelings after ECT treatment in responders (compared to before ECT). Negative bias is a known depressive symptom where maintained expectations about negative events often lead to unsurprising emotional reactions when such events arise, often contributing to flatter affect in patients with depression^64–66^. As such, ECT may alter cognitive-emotional processes, previously shown to be tracked by the inferior frontal operculum^67^, translating to increased emotional reactivity to punishments in responders.

The VPRL framework^16–18^ used here allowed us to identify potential computational phenotypes of ECT responders under hypotheses of independent reward and punishment learning systems in the brain. We identified specific learning and affective mechanisms that accompanied depression improvement in responders, which provided an objective, translatable understanding of successful ECT mechanisms^51^. Importantly, we recently demonstrated that the VPRL model tracks sub-second fluctuations in dopaminergic concentrations in the human brain using the same task used in this study^17^. And prior work has suggested that counterfactual signals can have a significantly influence on sub-second dopamine signals in humans^68^. In our visit 1 (pre-ECT) work, we showed that ‘worse-than-expected’ punishment prediction errors accounted for differences in ‘pre-ECT’ patients with TRD; here, we show that these differences were normalized in ECT treatment responders and correlated these changes to increased BOLD activity in the inferior frontal operculum. These results are consistent with prior work that suggested that punishment learning processes may be altered in depression^9^ but extend these studies by suggesting specific neurocomputational mechanisms underlying ECT symptom improvement. More work is needed to clarify the specificity of our proposed computational phenotypes of ECT treatment responders and whether these tools may be used to augment clinical work.

### Limitations

Our sample sizes for ECT non-responder group were small (N=4) and we did not have enough statistical power to run separate analyses to compare of investigate effects in non-responders. We aimed to observe the natural course of the current standard of care for ECT, therefore, ECT sessions varied by patient due to individual treatment plans that the research team was not involved with; however, this variation likely contributed to the overall successful treatment outcomes. Given this variation in treatment, but uniformity of treatment outcomes, our results may point to a general neurobehavioral state that may be a target for successful treatment. More work is needed to clarify these observations and test these hypotheses.

### Conclusions

We estimated the magnitude of dynamic neurobehavioral changes in patients receiving ECT by using neurocomputational models of learning and affective processes fit to behavioral data from patients with TRD. This study demonstrates the utility of applying computational models to assess *dynamic* behavioral, neural, cognitive, and emotional processes that cannot not be captured by current clinical assessments. Future work developing computationally derived estimate of neurobehavioral signals relevant to clinical decisions and patient behaviors is needed to further advance objective and quantitative tools into psychiatry where subjective reports and trial-and-error methods are the current standard of care.

## Supporting information

Supplementary Content

## Data Availability

All data produced in the present study are available upon reasonable request to

## Author Contributions

Kishida and Jones had full access to all the data in the study and take responsibility for the integrity of the data and the accuracy of the data analysis.

*Study concept and design:* Kishida, Gligorovic, Douglas, Jones, Sands.

*Acquisition, analysis, or interpretation of data:* All authors.

*Drafting of the manuscript:* Jones, Kishida.

*Critical revision of the manuscript for important intellectual content:* All authors.

*Statistical analysis:* Jones, Sands, Trattner.

*Obtained funding:* Kishida.

*Administrative, technical, or material support:* All authors.

*Study supervision:* Kishida, Gligorovic, Douglas, Ramos, Jones.

## Additional Contributions

We thank all of the individuals who took part in this study, clinic staff who assisted with recruitment, and MRI staff who assisted with research visits. We thank Dr. Mary Moya-Mendez for her work in advertisement and IRB development. The following research assistants from the Translational Neuroscience Department at Wake Forest School of Medicine were involved in data collection: Ashley Shipp, BS and Michael Howze, BS.

