## Supplementary Content for "Computational Phenotyping of Electroconvulsive Therapy Outcomes in Treatment-Resistant Depression": Final visit 2 Supplementary Online Content.docx

**eMethods.**

**eReferences.**

**eFigure 1.** Probabilistic reward and punishment with subjective rating (PRPwSR) task structure

**eFigure 2.** Behavioral performance across research visits for ECT, non-ECT, and no-depression groups

**eTable 1.** Variation of ECT treatment among patient groups

**eTable 2.** Participant demographic information

**eTable 3.** Current psychiatric medications and comorbidities reported by all participants with depression

**eTable 4**. Cohort differences in clinical scores across research visits

**eTable 5.** F-test for visit 2 – visit 1 changes in learning and subjective experience neurocomputations in ECT, non-ECT, and no-depression cohorts

**eMethods.**

**Study Design**

This study involved two research visits corresponding with the standard-of-care ECT treatment timeline (Fig.1A). Here, we report findings from “post-ECT” research “visit 2.” We recruited three participant groups according to depression diagnosis and ECT treatment plan (ECT depression, non-ECT depression, and no-depression groups). All participants completed a Probabilistic Reward and Punishment with Subjective Rating task while undergoing fMRI scanning. After scanning, participants completed clinical assessments, including the Patient Health Questionnaire 9 (PHQ-9)^1^, Hamilton Depression Rating Scale (HAM-D)^2^, and Montreal Cognitive Assessment (MOCA)^3^.

Research visits 1 and 2 were separated by the time it took ECT patients to complete their standard-of-care ECT series. Patients completed their first research visit approximately one week prior to starting ECT and completed their second research visit approximately one week after their ECT treatment concluded. The standard-of-care ECT treatment timeline included 5-12 ECT procedures over 2-3 weeks in which treatment concluded per clinical evaluation from each patients’ psychiatrist (see eTable1 for information on patient ECT treatment durations). Non-ECT participants with depression and no-depression participants completed both research visits according to the same ECT treatment timeline without intervention. As such, ECT, non-ECT , and no-depression participants completed their second research visit approximately one to two months after research visit 1.

**Participants**

All participants provided written informed consent and this study was approved by the Wake Forest University School of Medicine Institutional Review Board (IRB00056131). Inclusion criteria for all participants included age between 18 and 85 years. Exclusion criteria included individuals unable to provide written consent and verbal assent, individuals unable to understand task instructions or consent documents, and women who were pregnant specific to the MRI portion of the study.

We recruited three cohorts of participants. “ECT” patients included patients with treatment-resistant depression who consented to receive ECT for the first time at their non-research standard-of-care Atrium Health Wake Forest Baptist Outpatient Psychiatry and Behavioral Health visit (ECT, N=21; 47.6% female). Participants with (non-ECT, N=36; 69.4% female) and without depression (no-depression, N=38; 65.8% female) were recruited through advertisements in Winston-Salem, North Carolina. Non-ECT participants reported a current diagnosis of depression on the study’s screening form or scored mild or greater levels of depression severity on PHQ-9 and HAM-D assessments completed during research visits. No-depression participants reported no current diagnosis of depression on the study’s screening form and below threshold levels of depression severity on PHQ-9 and HAM-D assessments. Non-ECT and no-depression participants reported no prior history of ECT and did not receive ECT during the study period. We re-assigned two individuals who initially reported no current depression diagnosis from the no-depression group to the non-ECT depression group due to mild to moderate depression severity scores on both PHQ-9 and HAM-D measures reported during the study. Approximately half (n = 22) of non-ECT depression participants indicated having tried two or more different medications in the past to treat their depression, meeting some criteria for treatment-resistant depression. However, participants in the non-ECT depression group indicated never receiving ECT in the past and did not receive ECT during the study period.

We defined ECT treatment responders as patients who showed any clinical improvement following their standard-of-care ECT treatment series. Clinical measures included clinician-reported notes about patient responses and PHQ-9 and HAM-D score change following ECT treatment at research visits. If any clinical measure indicated depression improvement, a patient was considered an ECT responder. These criteria resulted in 17 ECT responders and 4 ECT non-responders.

Our visit 1 analyses included 29 pre-ECT, 40 non-ECT, and 41 no-depression participants. Across the two visits, four subjects withdrew, six subjects became non-compliant, and five subjects were lost to follow-up, totaling 21 ECT, 36 non-ECT, and 38 no-depression participants for visit 2 analyses. Participants without visit 1 imaging analyses (due to scanning issues) were also not included in visit 2 imaging analyses to ensure direct participant comparisons. Two participants did not undergo MRI at visit 2 due to contraindications, totaling 19 ECT, 35 non-ECT, and 38 no-depression participants for imaging analyses. See eTable2 for details about participant demographic information and eTable3 for information on depression participant psychiatric medications and comorbidities.

**Probabilistic Reward and Punishment with Subjective Rating (PRPwSR) Task**

The PRPwSR task included 150 trials with three phases of 25, 50, and 75 trials respectively (Fig.1B). On each trial, participants chose between one of two icons that were randomly paired. Chosen icons were reinforced probabilistically with either a monetary gain/no gain or loss/no loss. The task consisted of six total icons, including three win icons associated with 25%, 50%, or 75% chances of monetary gain (versus no money) and three loss icons associated with the same fixed probabilities of monetary loss (versus no money). To ensure participants did not remember icon values from their first research visit, a new set of icons were randomly assigned to each participant at research visit 2.

In phase 1, two of the three possible win icons were presented on each trial and the chosen icon was reinforced probabilistically by a reward of $1 or $0. Phase 2 introduced the three loss icons with the chosen icon reinforced probabilistically by a loss of $1 or $0. On each trial in phase 2, participants viewed pairs of win icons or loss icons only. In phase 3, the expected value of each win and loss icon changed in that icon probabilities stayed the same, but the dollar amounts presented changed. Here, pairs of win icons, loss icons, or win and loss icons could appear on a given trial.

After each trial in the PRPwSR task, there was a one-third chance of a rating screen appearing that asked participants “How do you feel about the last outcome?”. Participants reported their subjective feelings on the rating scale which ranged from “very bad” to “very good” and the next trial began. See eFig.1 for further details about the PRPwSR task structure and eFig.2 for participant performance measures.

**Computational Modeling**

We used hierarchical Bayesian analysis^4^ to fit the Valence Partitioned Reinforcement Learning (VPRL)^5–7^ computational model to participants’ choice behavior on the PRPwSR task (Fig.1B). Choice behavior was modeled strictly as a learning problem in a manner consistent with standard RL methods. Expected values and outcome prediction errors derived specifically from the VPRL framework were then used to fit a linear regression model to predict subjective ratings^6^ on those trials where participants were asked ‘how they felt’ about the outcome they had just experienced. Each individual was then assumed to be characterized by their unique set of behaviorally derived parameters, which we refer to as their *computational phenotype*^8^.

*Valence Partitioned Reinforcement Learning model*

The VPRL framework describes the specific hypotheses that independent positive and negative valence systems track expectations about rewarding and/or punishing contexts^5–7^. This approach was previously demonstrated to be a better fit to behavior on the PRPwSR task (compared to traditional univalent temporal difference RL models) and was shown to better explain sub-second fluctuations in extracellular dopamine levels in humans performing the PRPwSR task^7^.

In VPRL, temporal difference based Q-learning and prediction error models are used in parallel for the positive system (Eq. 1 and 2) and negative system (Eq. 3 and 4).

$$Q_{s_{t,}a_{t}}^{P}\leftarrow Q_{s_{t,}a_{t}}^{P}+\alpha^{P}\cdot\delta_{t}^{P}$$

*(1)*

$$\delta_{t}^{P}=\left\{ \begin{aligned} {outcome}_{t}+\gamma^{P}maxQ_{s_{t+1,}\tilde{\alpha}}^{P}-Q_{s_{t,}a_{t}}^{P} if {outcome}_{t}>0 \\ 0 +\gamma^{P}maxQ_{s_{t+1,}\tilde{\alpha}}^{P}-Q_{s_{t,}a_{t}}^{P} if {outcome}_{t}\leq0 \end{aligned} \right.$$

*(2)*

$$Q_{s_{t,}a_{t}}^{N}\leftarrow Q_{s_{t,}a_{t}}^{N}+\alpha^{N}\cdot\delta_{t}^{N}$$

*(3)*

$$\delta_{t}^{N}=\left\{ \begin{aligned} {|outcome}_{t}|+\gamma^{N}maxQ_{s_{t+1,}\tilde{\alpha}}^{N}-Q_{s_{t,}a_{t}}^{N} if {outcome}_{t}<0 \\ 0 +\gamma^{N}maxQ_{s_{t+1,}\tilde{\alpha}}^{N}-Q_{s_{t,}a_{t}}^{N} if {outcome}_{t}\geq0 \end{aligned} \right.$$

*(4)*

The superscripts *P* and *N* are used to denote the positive and negative systems, respectively. In Eq. 1, the positive system’s value estimate $Q_{s_{t,}a_{t}}^{P}$, of the quality, $Q$, of an action, $a_{t}$, in a given state, $s_{t}$, at time, $t$, is updated by the positive system’s reward prediction error, $\delta_{t}^{P}$. $\alpha^{P}$ is the positive system’s learning rate that weights the influence of $\delta_{t}^{P}$ updates of the value estimate, $Q_{s_{t,}a_{t}}^{P}.$In Eq. 2, ${outcome}_{t}$ is the reward experienced at time *t*. The discounting factor ($\gamma^{P}$) weighs the extent that the value of the future is considered; $maxQ_{s_{t+1,}\tilde{\alpha}}^{P}$ represents the value of the future state after choosing action $\alpha$ from the available actions ($\tilde{\alpha}$) that maximize the expected value ($maxQ^{P}$) in the future state ($s_{t+1}$). Eq. 3 and 4 describe the negative system’s value estimates ($Q_{s_{t,}a_{t}}^{N}$) and punishment prediction errors ($\delta_{t}^{N}$).

The overall estimate of value is obtained by integrating the independently estimated positive and negative system Q-values (Eq. 5):

$$Q_{s_{t},a_{t}}= Q_{s_{t,}a_{t}}^{P}- Q_{s_{t,}a_{t}}^{N}$$

*(5)*

Participants’ overall estimated Q-values were inserted into a softmax policy function (Eq. 6) to estimate the probabilities of choosing icons when presented in the PRPwSR task:

$$P\left( choice={choice}_{1} \right| Q_{s_{t},{choice}_{1}}, Q_{s_{t},{choice}_{2}})= \frac{e^{Q_{s_{t},{choice}_{1}}/\tau}}{e^{Q_{s_{t},{choice}_{1}}/\tau}+ e^{Q_{s_{t},{choice}_{2}}/\tau}}$$

*(6)*

Here, “$\tau$” is the choice temperature parameter constrained to a range of 0-20 that captures how deterministically ($\tau$ closer to 0) versus randomly ($\tau$ closer to 1) participants distribute their choices given their current value estimates.

For each of the cohorts (ECT, ECT responder, ECT non-responder, non-ECT, and no-depression), we fit the same VPRL model using hierarchical Bayesian analysis with uniform priors to derive parameter estimates pooled toward the respective group mean^4^. See Sands et al. for a mathematical description of the hierarchical Bayesian methods that we used here^6,7^.

We ran Hamiltonian Monte Carlo with No-U-Turn Sampler (HMC-NUTS) via Stan’s rstan interface to estimate the posterior distributions of VPRL model parameters across cohort data (version 4.2.2)^9^. For each cohort model fit, we ran four Markov chains, with 12,000 total samples per chain (8,000 after discarding warm-up samples). In addition, we confirmed that all Gelman-Rubin $\hat{R}$ values were approximately 1 for all parameters indicating good chain mixing.

*Subjective Feeling regression model*

We used Bayesian linear regression^10^ to estimate participant’s momentary feelings ratings throughout the PRPwSR task using VPRL-model-derived Q-values ($\boldsymbol{Q}_{\boldsymbol{s}_{\boldsymbol{t,}}\boldsymbol{a}_{\boldsymbol{t,}}\boldsymbol{chosen}}^{\boldsymbol{P}}\boldsymbol{,}\boldsymbol{Q}_{\boldsymbol{s}_{\boldsymbol{t,}}\boldsymbol{a}_{\boldsymbol{t,}}\boldsymbol{unchosen}}^{\boldsymbol{P}}\boldsymbol{,}\boldsymbol{Q}_{\boldsymbol{s}_{\boldsymbol{t,}}\boldsymbol{a}_{\boldsymbol{t,}}\boldsymbol{chosen}}^{\boldsymbol{N}}\boldsymbol{,}\boldsymbol{Q}_{\boldsymbol{s}_{\boldsymbol{t,}}\boldsymbol{a}_{\boldsymbol{t,}}\boldsymbol{unchosen}}^{\boldsymbol{N}})$and outcome prediction errors (**+**$\boldsymbol{\delta}_{\boldsymbol{t}}^{\boldsymbol{P}}$, **-**$\boldsymbol{\delta}_{\boldsymbol{t}}^{\boldsymbol{P}}$, **+**$\boldsymbol{\delta}_{\boldsymbol{t}}^{\boldsymbol{N}}$, **-**$\boldsymbol{\delta}_{\boldsymbol{t}}^{\boldsymbol{N}}$) experienced during the 50 rating trials as the independent variables and the subjective rating as the dependent variable. The positive and negative sign on these error signals indicated greater-than (+) or less-than (-) expected and are separated for these models to reflect the hypothesis that these are not symmetrically weighted in their influence on affective responses.

The Subjective Feeling regression model was fit using Bayesian methods:

$$E\left( D_{i} | \beta,X \right)=\beta_{0}+\beta_{1}x_{1}+\beta_{2}x_{2}+\beta_{3}x_{3}+\beta_{4}x_{4}+\beta_{5}x_{5}+\beta_{6}x_{6}+\beta_{7}x_{7}+\beta_{8}x_{8}+\varepsilon_{i}$$

*(7)*

$$\varepsilon\sim Normal(0,\sigma^{2})$$

The expected value of a rating on trial *i* is denoted $E(D_{i}|\beta,X)$ and is assumed to be normally distributed. The mean of the subjective rating on a trial ($D_{i}$) is a linear function of positive and negative system prediction errors and expected value of chosen and unchosen option predictor matrices ($X)$, with coefficients of the linear combination represented by $\beta$ vector. The predictor variables include Q-values of chosen and unchosen reward and loss icons ($Q_{s_{t,}a_{t,}chosen}^{P}, Q_{s_{t,}a_{t,}unchosen}^{P}, Q_{s_{t,}a_{t,}chosen}^{N}, Q_{s_{t,}a_{t,}unchosen}^{N})$ and reward and punishment prediction errors (+$\delta_{t}^{P}$, -$\delta_{t}^{P}$, +$\delta_{t}^{N}$, -$\delta_{t}^{N}$) that occur during the 50 rating trials for each participant, totaling eight model predictors. $\varepsilon_{i}$ are the normally distributed errors with variance $\sigma^{2}.$

We fit the Bayesian regression model to each cohort and implemented a leave-one-out cross-validation approach where each left-out participant’s posterior distributions of model coefficients were informed by their respective cohort (i.e., group informed). The Bayesian regression model generated posterior distributions of coefficient values for each of the eight independent learning variables as well as the constant $\beta_{0}$term. See Sands et al. for a mathematical description of the Bayesian methods used here^6^.

For each cohort model fit, we excluded 1,000 warm-up samples which produced four parallel chains of length 2,500 for a total of 10,000 samples for each model parameter. $\hat{R}$ values were approximately 1 for all parameters.

**Functional data acquisition and MRI pre-processing**

*Data acquisition*

We utilized a multi-band echo-planar imaging (EPI) sequence to record BOLD data during the PRPwSR task using a Siemens MAGNETOM 3T Skyra whole-body scanner with a 32-channel head coil (MB factor = 8; TR = 1000ms; TE = 30ms; flip angle = 52 degrees; FOV = 20.8 cm; 72 interleaved sagittal slices; isotropic 2mm3 voxels). We also acquired a high-resolution T1-weighted anatomical scan.

*Pre-processing*

We performed all data pre-processing using FSL and SPM12^11,12^. This included first estimating head motion via registration to a single-band reference image (SBRef); correcting for EPI (B0) distortion via a fieldmap estimated using reverse-phase encoded functional volumes in the R-L and L-R directions through FSL’s topup tool^13^; co-registering volumes to the 0.5x0.5x1 mm^3^ T1-weighted structural image and then to the MNI template space; spatially smoothing with a 4mm FWHM Gaussian filter; high-pass filtering at 128sec (<0.008Hz); and normalizing by the session grand-mean value. For two non-ECT participants, we utilized FSL’s FUGUE to correct for EPI distortions from acquired fieldmaps due to scanning issues obtaining the reverse-phase encoded functional volumes.

*Model-based analyses*

For each participant, we fit two first-level general linear models (GLMs) to model blood-oxygen-level-dependent (BOLD) activity that occurred during PRPwSR task events. Regressors of interest were convolved with a canonical hemodynamic response function. The first GLM included the option presentation timepoint parametrically modulated by participants’ icon expected values ($Q_{s_{t,}a_{t,}chosen}^{P}, Q_{s_{t,}a_{t,}unchosen}^{P}, Q_{s_{t,}a_{t,}chosen}^{N}, Q_{s_{t,}a_{t,}unchosen}^{N})$and the outcome presentation timepoint parametrically modulated by participants’ prediction errors (+$\delta_{t}^{P}$, -$\delta_{t}^{P}$, +$\delta_{t}^{N}$, -$\delta_{t}^{N}$) across all task trials. This allowed us to model learning neurocomputations. The second GLM included the outcome presentation timepoint parametrically modulated by participants’ expected values and prediction errors weighted by their respective (median) Subjective Feeling model coefficients (e.g., +$\delta_{t}^{P}\cdot\beta_{+\delta_{t}^{P}}$). This allowed us to model affective neurocomputations. For the GLMs, regressors were derived from individual-level parameter estimates from VPRL and Subjective Feeling models and all regressors were z-scored. All motor and visual task measures and six head motion parameters were also included as regressors of no interest.

Based on our behavioral results, we built visit 2 – visit 1 contrasts for regressors of interest. We performed second-level whole-brain one-sample t-tests in ECT responders for contrasts of interest to assess BOLD activity changes across research visits. We then performed second-level whole-brain analysis of the variance (ANOVA) on contrasts of interest for ECT, non-ECT, and no-depression groups. All contrasts assessed were based on observed behavioral changes (Table1A-B). All statistical analyses were conducted at an uncorrected threshold of p<0.001 and reported results were selected using a family-wise error (FWE)-corrected threshold of p<0.05 at cluster and peak voxel levels.

**Statistical Analyses**

Hierarchical Bayesian analyses^4^ were performed to estimate posterior distributions of free parameters in the VPRL models of choice behavior and learning ($\boldsymbol{\alpha}^{\boldsymbol{P}}$,$\boldsymbol{\alpha}^{\boldsymbol{N}}$**,** $\boldsymbol{\gamma}^{\boldsymbol{P}}$, $\boldsymbol{\gamma}^{\boldsymbol{N}}$, and $\boldsymbol{\tau}$) and subsequently for coefficient parameters in the Subjective Feeling linear regression model ($\boldsymbol{\beta}_{\boldsymbol{constant}}\boldsymbol{,}\boldsymbol{\beta}_{\mathbf{+}\boldsymbol{\delta}_{\boldsymbol{t}}^{\boldsymbol{P}}}\boldsymbol{,}\boldsymbol{\beta}_{\mathbf{-}\boldsymbol{\delta}_{\boldsymbol{t}}^{\boldsymbol{P}}}\boldsymbol{,}\boldsymbol{\beta}_{\mathbf{+}\boldsymbol{\delta}_{\boldsymbol{t}}^{\boldsymbol{N}}}\boldsymbol{,}\boldsymbol{\beta}_{\boldsymbol{-\delta}_{\boldsymbol{t}}^{\boldsymbol{N}}}\boldsymbol{,}\boldsymbol{\beta}_{\boldsymbol{Q}_{\boldsymbol{s}_{\boldsymbol{t,}}\boldsymbol{a}_{\boldsymbol{t,}}\boldsymbol{chosen}}^{\boldsymbol{P}}}\boldsymbol{,}\boldsymbol{\beta}_{\boldsymbol{Q}_{\boldsymbol{s}_{\boldsymbol{t,}}\boldsymbol{a}_{\boldsymbol{t,}}\boldsymbol{unchosen}}^{\boldsymbol{P}}}\boldsymbol{,}\boldsymbol{\beta}_{\boldsymbol{Q}_{\boldsymbol{s}_{\boldsymbol{t,}}\boldsymbol{a}_{\boldsymbol{t,}}\boldsymbol{chosen}}^{\boldsymbol{N}}}\boldsymbol{,}\boldsymbol{\beta}_{\boldsymbol{Q}_{\boldsymbol{s}_{\boldsymbol{t,}}\boldsymbol{a}_{\boldsymbol{t,}}\boldsymbol{unchosen}}^{\boldsymbol{N}}}$). Cohort posterior distribution differences were determined by examining 95% highest density intervals (HDI) that excluded 0 or that were within 0.05 of excluding 0^14^ (while this threshold is arbitrary, this acknowledges the limitations of overly strict significance thresholds and allowed us to consider valuable information that may have been missed with stricter cutoffs).

**eFigure 1. Probabilistic reward and punishment with subjective rating (PRPwSR) task structure**

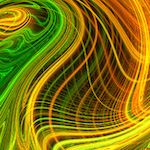

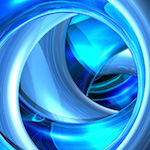

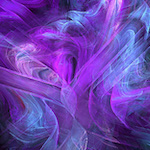

Trial

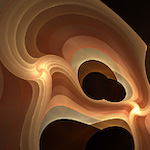

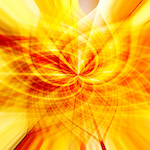

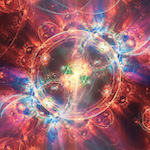

1

25

75

150

0.25(-$1.00) = -$0.25

0.50(-$1.00) = -$0.50

0.75($1.00) = -$0.75

0.25($1.00) = $0.25

0.50($1.00) = $0.50

0.75($1.00) = $0.75

0.75($0.50) = $0.375

0.25($2.50) = $0.625

0.50($1.50) = $0.75

0.25(-$1.25) = -$0.3125

0.50(-$0.75) = -$0.375

0.75(-$1.00) = -$0.1875

Icon value =

outcome probability ($)

**Phase I**

**Phase II**

**Phase III**

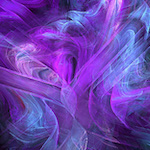

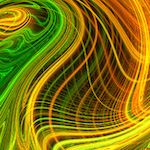

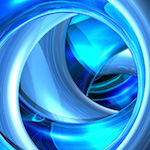

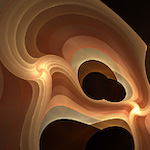

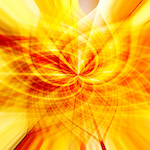

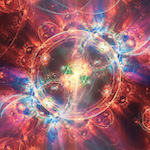

Win Icons

Loss Icons

Depicted is an example of six fractal images presented throughout the PRPwSR task. Icon expected values are the product of the icon probability and dollar amount presented (e.g., purple icon in phase 1: 0.75 probability of $1 = $0.75 value). Phase 1 includes 3 win icons with fixed probabilities of earning $1 (versus $0). Phase 2 introduces three loss icons with fixed probabilities of losing $1 (versus $0). In phase 3, icon probabilities remain the same, but the dollar amounts they present change. This changes icon expected values (e.g., purple icon in phase 1 and 2 displays $1 but in phase 3 displays $0.50; expected value transitions from $0.75 to $0.37).

**eFigure 2. Behavioral performance across research visits for ECT, non-ECT, and no-depression groups**

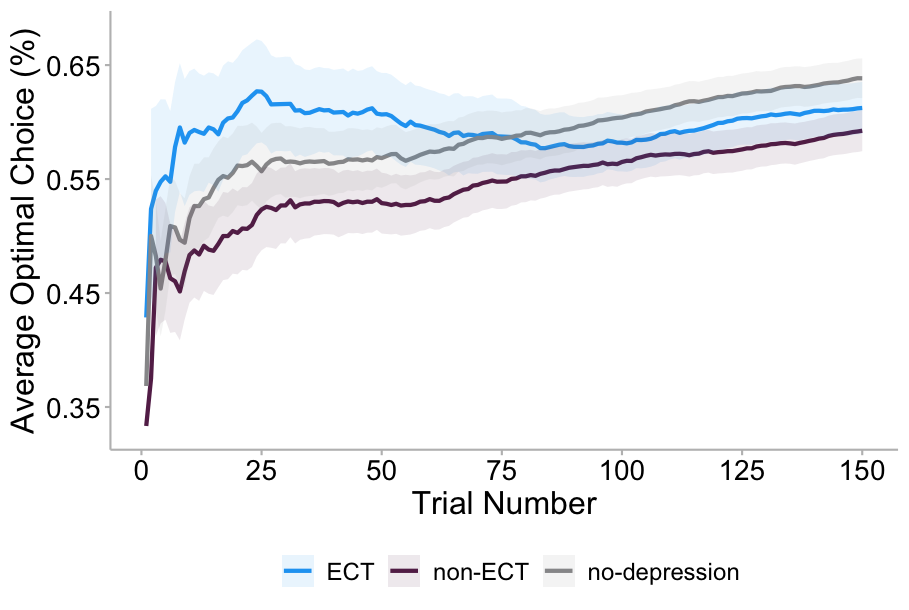

**A**

**B**

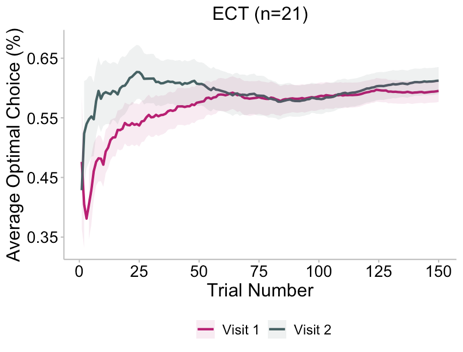

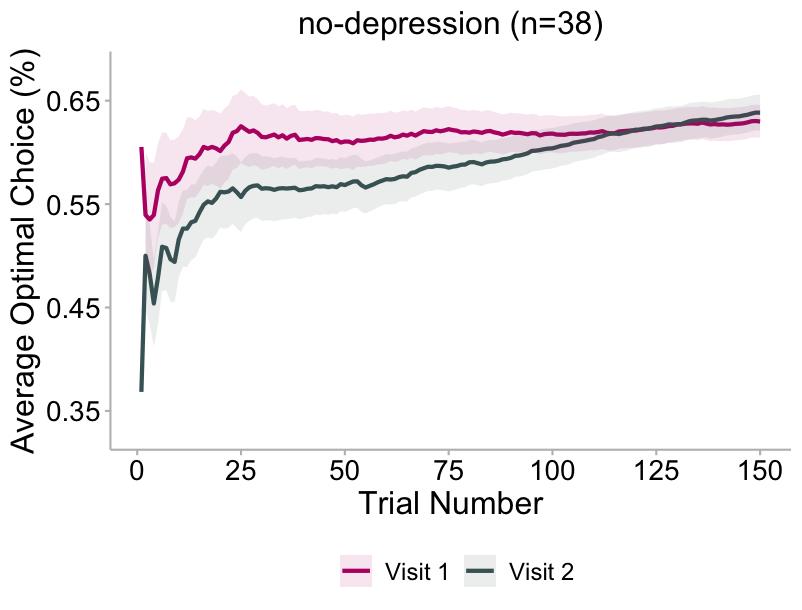

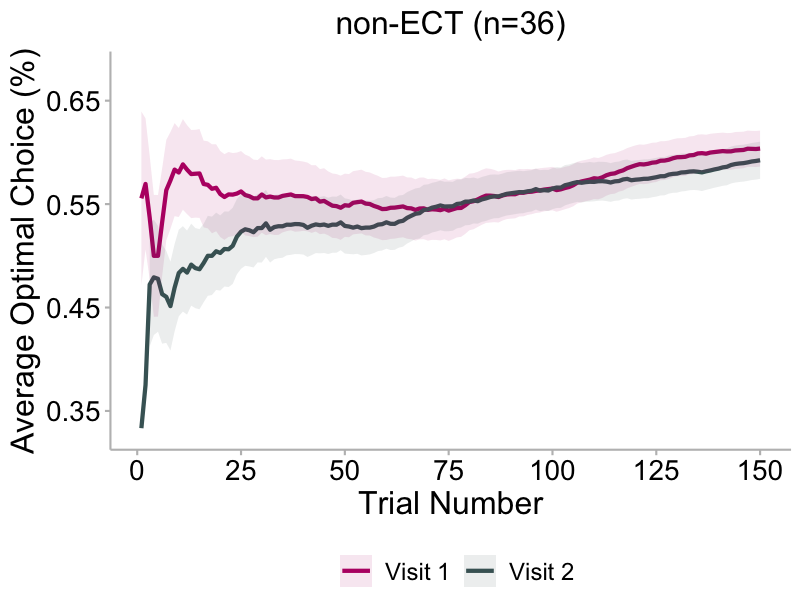

Trial-by-trail descriptions of the average optimal choices for each cohort on the PRPwSR task. Optimal choices indicate selection of the icon with the highest monetary reward value (for win icons) and lowest monetary loss value (for loss icons). **A)** ECT (blue), non-ECT (purple), and no-depression (gray) cohort mean optimal choices across all 150 trials in research visit 2. **B)** Mean optimal choices across trials for visit 1 (pink) and visit 2 (dark green) for ECT (left), non-ECT (middle), and no-depression (right) cohorts. Solid lines are shown with 95% confidence intervals.

| **eTable 1.** **Variation of ECT treatment among patient groups** | | |
| --- | --- | --- |
| Group | *# of ECT procedures*  *Mean [SD]; Range* | *Duration of procedures (days)*  *Mean [SD]; Range* |
| All ECT patients (N=21) | 9.29 [3.64]; 3-20 | 30.86 [14.17]; 2-62 |
| ECT Responders (N=17) | 9.29 [3.65]; 3-20 | 31.18 [14.45]; 2-62 |
| ECT Non-responders (N=4) | 9.25 [4.11]; 4-14 | 29.50 [14.93]; 9-42 |

| **eTable 2. Participant demographic information** | | | | |
| --- | --- | --- | --- | --- |
| Demographic Variable | ECT  (n = 21) | Non-ECT  (n = 36) | No-depression  (n = 38) | Group Comparison |
| Age, mean (SD), years | 40.19 (15.60) | 41.17 (15.23) | 40.71 (15.08) | F_2,92_ = 0.028 (ns) |
| Gender |  |  |  | $\chi_{4}^{2}=$ 5.72 (ns) |
| Male | 10 | 11 | 13 |  |
| Female | 10 | 25 | 25 |  |
| Non-binary | 1 | 0 | 0 |  |
| Race |  |  |  | $\chi_{8}^{2}=$ 9.69 (ns) |
| AIAN | 1 | 0 | 0 |  |
| Asian | 0 | 1 | 4 |  |
| Black/African American | 3 | 3 | 5 |  |
| White/Caucasian | 17 | 32 | 29 |  |
| Other | 1 | 0 | 1 |  |
| Ethnicity |  |  |  | $\chi_{2}^{2}=$ 3.30 (ns) |
| Hispanic/Latino | 2 | 2 | 0 |  |
| Not Hispanic/Latino | 19 | 34 | 38 |  |
| Abbreviations: ns: not significant; AIAN: American Indian and Alaska Native. One subject reported multiple races (data reported as count) where a multirace category was included in significance testing. | | | | |

| **eTable 3.** **Current psychiatric medications and comorbidities reported by all participants with depression** | | |
| --- | --- | --- |
|  | ECT  (n = 21) | Non-ECT  (n = 36) |
| Medications |  |  |
| Antidepressants^a^ | 15 | 25 |
| Antipsychotic/Anticonvulsant/Mood Stabilizer | 12 | 9 |
| Anxiolytic/Hypnotic^b^ | 4 | 9 |
| Stimulant | 2 | 4 |
| None reported | 1 | 7 |
| Comorbidities |  |  |
| Anxiety/Panic Disorder | 14 | 17 |
| Attention-Deficit/Hyperactivity Disorder | 4 | 5 |
| Bipolar Disorder | 5 | 4 |
| Borderline or Dissociative Personality Disorder | 4 | 1 |
| Eating Disorder | 2 | 1 |
| Obsessive Compulsive Disorder | 0 | 2 |
| Post Traumatic Stress Disorder | 5 | 1 |
| Psychosis/Schizophrenia/Schizoaffective Disorder | 0 | 1 |
| Sleep Disorder | 2 | 0 |
| None reported | 4 | 14 |
| ^a^Antidepressants included SSRIs, SNRIs, NDRIs, SARIs, and TCAs. ^b^Anxiolytics/hypnotics included included benzodiazepines and antihistamines. | | |

| **eTable 4**. **Cohort differences in clinical scores across research visits** | | | | | | | | | | | |
| --- | --- | --- | --- | --- | --- | --- | --- | --- | --- | --- | --- |
| *Variable* | *ECT (n = 21)* | | | *Non-ECT (n = 36)* | | | *No-depression (n = 38)* | | | *Group Comparisons of Clinical Changes Between Visits* | |
|  | *pre-ECT* | *post-ECT* | *t(20)* | *Visit 1* | *Visit 2* | *t(35)* | *Visit 1* | *Visit 2* | *t(37)* | *F(2,92)* | *Post-hoc* |
| *MOCA*  *Mean (SD)* | 26.95 (2.62) | 26.14 (2.65) | -1.62 | 26.29 (3.45) | 26.71 (4.03) | 0.80 | 26.97 (2.05) | 28.37 (1.81) | **4.15^a^** | **4.35^c^** | **ECT-No-depression^c^**  ECT-non-ECT  Non-ECT-No-depression |
| *PHQ-9*  *Mean (SD)* | 19.0 (5.80) | 11.95 (7.26) | **-4.10^a^** | 11.91 (6.58) | 8.94 (5.75) | **-3.29^b^** | 1.68 (2.58) | 1.74 (2.64) | 0.23 | **14.54^a^** | **ECT-No-depression^a^**  **ECT-non-ECT^b^**  **Non-ECT-No-depression^c^** |
| *HAM-D*  *Mean (SD)* | 24.76 (6.76) | 18.0 (9.53) | **-3.64^b^** | 14.12 (8.27) | 11.21 (8.37) | **-2.66^c^** | 1.97 (2.17) | 1.71 (2.10) | -1.03 | **8.34^a^** | **ECT-No-depression^a^**  ECT-non-ECT  Non-ECT-No-depression |
| Abbreviations: MOCA: Montreal Cognitive Assessment; PHQ-9: Patient Health Questionnaire-9; HAM-D: Hamilton Depression Rating Scale. Paired t-tests were performed for within-cohort comparisons. One-way ANOVA with tukey’s post-hoc testing was performed for between-cohort comparisons. ^a^p<0.001, ^b^p<0.01, ^c^p<0.05. | | | | | | | | | | | |

| **eTable 5. F-test for visit 2 - visit 1 changes in learning and subjective experience neurocomputations in ECT, non-ECT, and no-depression cohorts** | | | | | | |
| --- | --- | --- | --- | --- | --- | --- |
| **Parameter** | **Region** | **Voxels** | **Cluster-level p-value** | **Peak-level**  **p-value** | **Statistic** | **Peak MNI coordinates**  **[x y z]** |
| A. *Learning Computations* | | | | | | |
| ${\Delta_{visit 2-visit 1}(\beta}_{GLM}(\delta_{t}^{P}))$ |  |  |  |  | NS |  |
| B. *Subjective Experience Computations* | | | | | | |
| ${\Delta_{visit 2-visit 1}(\beta}_{GLM}(\beta_{Q_{s_{t,}a_{t,}unchosen}^{P}}))$ |  |  |  |  | NS |  |
| ${\Delta_{visit 2-visit 1}(\beta}_{GLM}(\beta_{+\delta_{t}^{N}}))$ |  |  |  |  | NS |  |
| ${\Delta_{visit 2-visit 1}(\beta}_{GLM}(\beta_{-\delta_{t}^{N}}))$ | Right calcarine gyrus | 65 | **0.028** | 0.467 | F(2,88) = 13.17 | [12 -70 6] |
|  | Right calcarine gyrus | 148 | **0.002** | 0.075 | $\dagger$non-ECT > ECT | [12 -70 6] |
|  | Right precuneus | 95 | **0.018** | 0.737 | $\dagger$non-ECT > ECT | [16 -62 40] |
|  | Left posterior cingulum | 111 | **0.009** | 0.904 | $\dagger$non-ECT > ECT | [-2 -38 24] |
|  | Right angular gyrus | 87 | **0.027** | 0.811 | $\dagger$no-depression > ECT | [36 -52 26] |
| All analyses were performed at an uncorrected threshold of p<0.001 in which reported results were selected using an FWE-corrected threshold of p<0.05 at cluster and peak voxel levels. Bold font indicates significance. NS: not significant. $\boldsymbol{\dagger:}$ post-hoc t-test. | | | | | | |
